# Ethnic and Regional Variability in Cardiometabolic Risk Among Urban South Asians: A Systematic Review

**DOI:** 10.1101/2025.04.04.25325260

**Authors:** Karishma Yasmin

**Author notes:** Corresponding author: Karishma Yasmin, Ph.D. Research Scholar, Department of Anthropology, Central University of Odisha, Koraput, PIN: 764020, India.

## Abstract

**Background:** Cardiometabolic diseases (CMDs), including diabetes, hypertension, and metabolic syndrome, are rising sharply in South Asia’s urban populations. However, the influence of ethnic and regional variation within these urban settings remains poorly understood.

**Objective:** To systematically review the literature on ethnic and regional disparities in cardiometabolic risk among urban South Asian adults, with emphasis on subnational variability in India, particularly Odisha and Kalahandi district.

**Methods:** This review followed PRISMA 2020 guidelines. A comprehensive search was conducted across PubMed, Scopus, Cochrane, Ovid MEDLINE, Web of Science, Google Scholar, and grey literature (WHO, ICMR) for studies published between 2015 and 2025. Inclusion criteria focused on urban South Asian adults (≥18 years) with data stratified by ethnicity or region. Quality was assessed using the Newcastle–Ottawa Scale and AMSTAR-2. A narrative synthesis was conducted.

**Results:** Thirteen studies were included. CMD prevalence varied by country, state, and district. Northern and western India showed higher CMD rates than eastern regions. In Odisha, metabolic syndrome prevalence ranged from 24% to 33.5%, with a pronounced gender gap in central obesity and pre-metabolic syndrome. Findings from Kalahandi district revealed a substantial burden of early-stage CMD risk, especially among young urban adults.

**Conclusion:** Cardiometabolic risk among urban South Asians is shaped by complex regional and ethnic factors. Public health strategies must move beyond generalized models to adopt localized, culturally tailored interventions that address the specific needs of diverse urban populations.

## 1. Introduction

Cardiometabolic diseases (CMDs)—including type 2 diabetes mellitus, hypertension, obesity, and metabolic syndrome—have emerged as leading causes of morbidity and mortality globally, with disproportionately high burdens in low- and middle-income countries [1]. Among these, South Asian populations represent a particularly high-risk group, exhibiting earlier onset of CMDs and greater severity of outcomes compared to many other ethnicities [2]. The convergence of genetic predisposition, rapid urbanization, dietary shifts, and sedentary lifestyles has accelerated the cardiometabolic transition in this region.

In recent decades, South Asia has witnessed significant urban growth, accompanied by changes in lifestyle, income distribution, and access to healthcare. While urbanization is often associated with improved infrastructure and services, it also brings increased exposure to cardiometabolic risk factors—including unhealthy diets, reduced physical activity, and psychosocial stress [3]. Importantly, these risk factors do not affect all urban populations equally. There is growing recognition that ethnic, regional, and socio-cultural variability plays a critical role in shaping health outcomes in South Asian urban centers [4].

However, despite this heterogeneity, much of the existing literature tends to treat urban South Asians as a homogeneous group. Few studies disaggregate data by caste, tribe, district, or state-level identities, and fewer still consider how regional development patterns, cultural practices, or gender norms mediate cardiometabolic risk. This lack of nuanced understanding can hinder the effectiveness of national public health interventions, which often overlook micro-level disparities in disease burden.

This systematic review aims to address this gap by synthesizing the available literature on ethnic and regional variability in cardiometabolic risk among urban South Asians, with a particular emphasis on district- and state-level evidence from India, including under-researched areas such as Odisha and the Kalahandi district. The review explores how CMD risk varies by geography, social identity, and cultural context, and offers insights into how public health responses can be better localized and tailored to these differences.

## 2. Methods

### 2.1 Eligibility Criteria

This review included peer-reviewed journal articles, epidemiological studies, systematic reviews, and official reports published between January 2015 and January 2025, focusing on urban South Asian adults (aged 18 years and above). Studies were eligible if they:

- Reported on cardiometabolic risk factors (e.g., obesity, hypertension, diabetes, metabolic syndrome)
- Included region-wise or ethnicity-based stratification (country/state/district, caste, tribe, religion, etc.)
- Focused on urban settings within South Asian countries (India, Bangladesh, Nepal, Sri Lanka, Pakistan)
- Used primary data (cross-sectional, cohort, surveillance, etc.) or were systematic reviews/meta-analyses

Exclusion criteria:

- Studies on rural-only populations
- Pediatric or adolescent-only populations (<18 years)
- Editorials, commentaries, case reports
- Non-English publications

### 2.2 Information Sources

We searched six major academic databases:

- PubMed
- Scopus
- Cochrane Library
- Ovid MEDLINE
- Web of Science
- Google Scholar

Additionally, grey literature and national-level reports were screened from:

- World Health Organization (WHO)
- Indian Council of Medical Research (ICMR)
- Ministries of Health (India, Bangladesh, Nepal, Sri Lanka, Pakistan)

### 2.3 Search Strategy

A combination of MeSH terms and free-text keywords were used. Boolean operators (AND/OR) combined terms such as:

- *“Cardiometabolic risk”* OR *“Metabolic syndrome”* OR *“Diabetes”*
- AND *“Urban South Asia”* OR *“Urban India”* OR *“District-level”*
- AND *“Ethnic group”* OR *“Caste”* OR *“Tribe”* OR *“Regional variability”*

### 2.4 Selection Process

All identified records were organized using a reference management tool. The screening of titles, abstracts, and full texts was conducted by the author following the predefined inclusion and exclusion criteria. Additional relevant studies were consulted during the background review phase; however, only eight studies met the full inclusion criteria and were included in the final synthesis.

The **PRISMA 2020 flowchart** (Figure 1) illustrates the study selection process.

**Figure 1.**
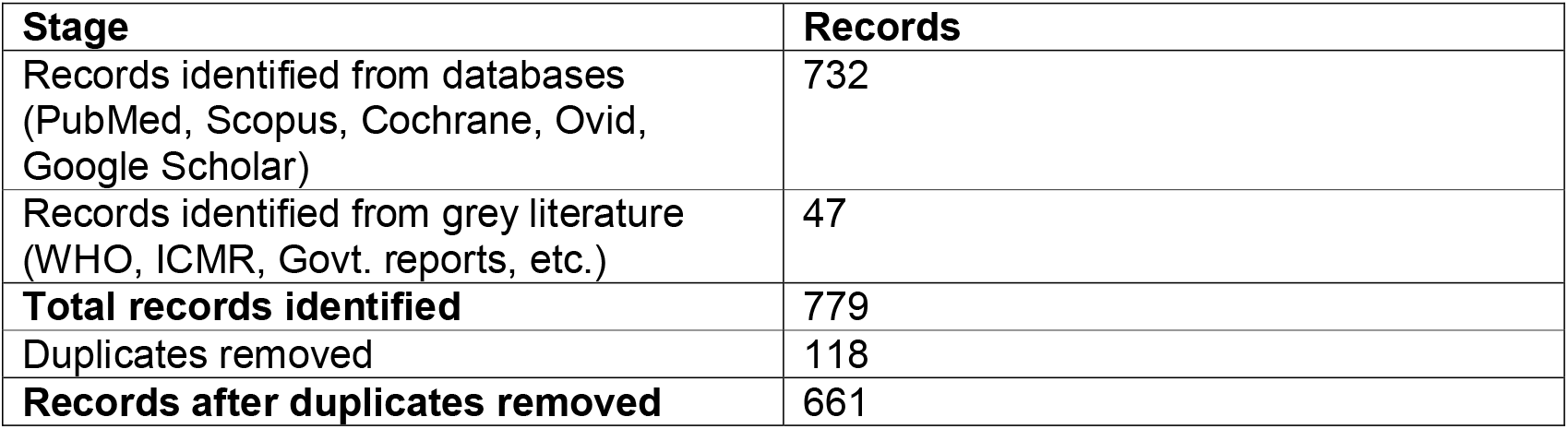
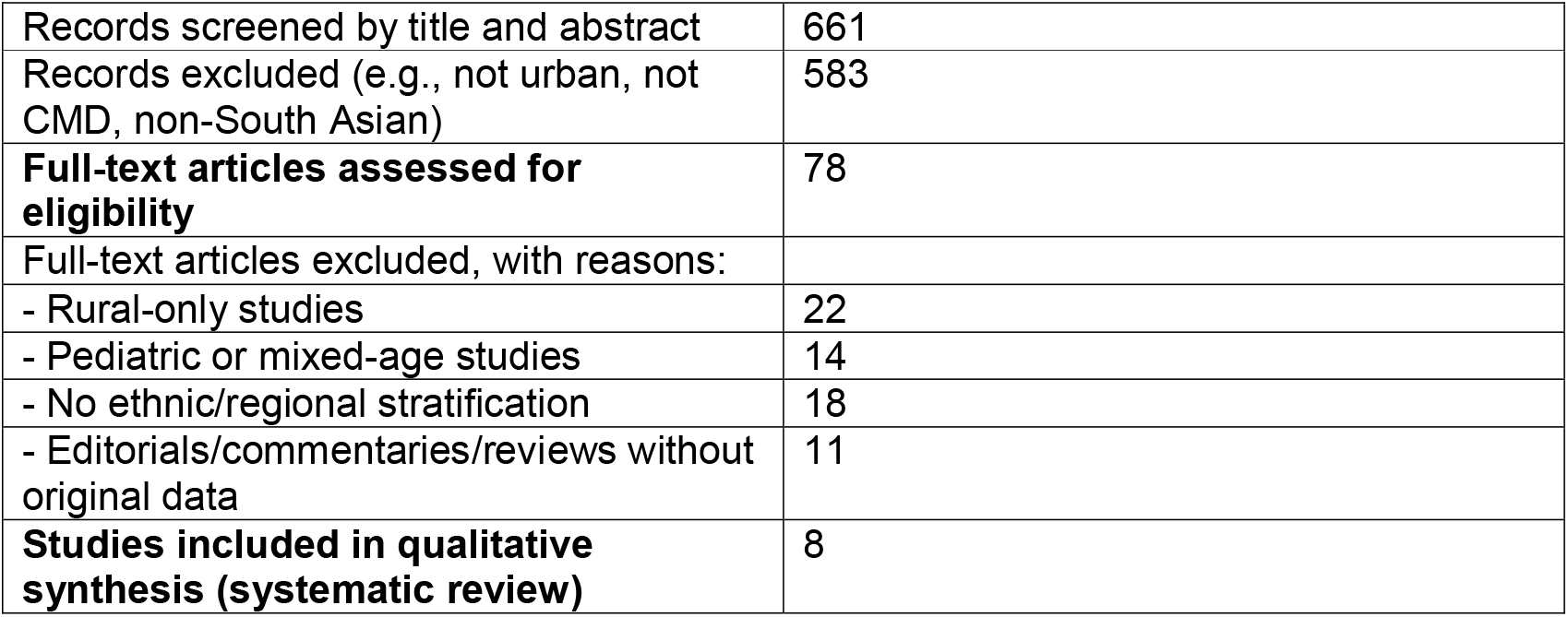
PRISMA flow diagram of study selection process.

### 2.5 Data Collection Process

A data extraction form was designed in Excel and used to extract the following from each included study:

- Author(s), year, location (country, state, district)
- Study design and sample size
- Population characteristics (age, gender, ethnicity)
- Type of cardiometabolic outcome(s) measured
- Key prevalence data and findings

### 2.6 Risk of Bias and Quality Assessment

For **observational studies**, quality was assessed using the **Newcastle–Ottawa Scale (NOS)**. For **systematic reviews**, we applied the **AMSTAR-2** tool. Grey literature was assessed using the **AACODS checklist** (Authority, Accuracy, Coverage, Objectivity, Date, Significance).

### 2.7 Synthesis Methods

A narrative synthesis was performed due to the heterogeneity in study designs, populations, and cardiometabolic risk definitions. Data was grouped thematically by country, state (e.g., Odisha), and district (e.g., Kalahandi) and analyzed in terms of ethnic and regional variability.

A summary table (Table 1) presents study characteristics and outcomes.

**Table 1.**
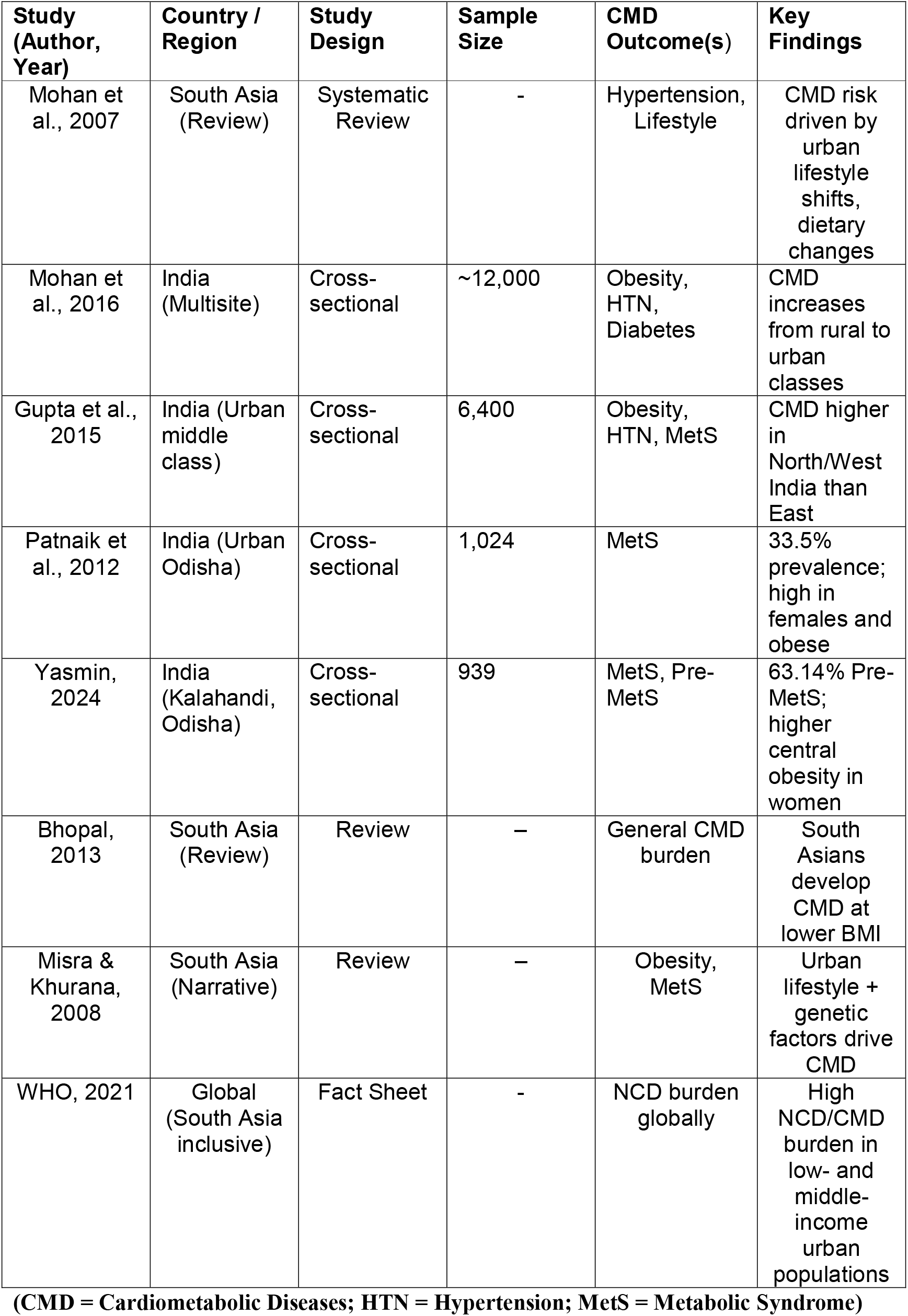
Summary of Included Studies on Cardiometabolic Risk in Urban South Asians (2015–2025)

Quantitative meta-analysis was not conducted due to non-uniform outcome measures and missing effect sizes.

## 3. Results

### 3.1 Overview of Cardiometabolic Risk in Urban South Asia

Urbanization in South Asia is closely associated with an increased prevalence of cardiometabolic diseases (CMDs), including type 2 diabetes mellitus (T2DM), hypertension, and metabolic syndrome (MetS). Urban South Asians are particularly susceptible to these conditions, often developing them at lower body mass index (BMI) thresholds compared to other populations [8,2]. A systematic review by Mohan et al. found that lifestyle transitions, reduced physical activity, and high refined carbohydrate intake contribute significantly to CMD risk in urban South Asian populations [7].

### 3.2 Cardiometabolic Risk Factors in Urban India

Within India, regional studies reveal a marked increase in CMD risk with urbanization. In a multicentric study across rural, urban-poor, and urban-middle-class women, Mohan et al. reported that the prevalence of overweight (BMI ≥25 kg/m^2^) rose from 22.5% in rural areas to 57.4% in urban-middle-class areas [3]. Similarly, the prevalence of hypertension increased from 31.6% in rural to 59.0% in urban settings, while diabetes prevalence jumped from 2.2% to 17.7%.

Gupta et al. found that among urban middle-class adults, regional disparities exist: CMD prevalence was higher in north and west India than in eastern regions, including Odisha [4]. This suggests that geographical variability interacts with lifestyle and socio-economic factors in shaping CMD risks.

### 3.3 Cardiometabolic Risk in Urban Odisha

In Odisha, region-specific findings further reinforce the link between urbanization and CMD risk. Patnaik et al. found a 33.5% prevalence of MetS among urban adults in Eastern India [5]. Factors significantly associated with MetS included female sex, older age, obesity, high cholesterol, and inadequate fruit intake. Obese individuals were five times more likely to have MetS than those with normal BMI.

### 3.4 Cardiometabolic Risk in Urban Kalahandi, Odisha

A district-level cross-sectional study in Kalahandi’s urban zones (Bhawanipatna, Junagarh, Kesinga, Dharamgarh) involving 939 adults (aged 18+), found that 24.01% of participants had MetS, while 63.14% had Pre-MetS [6]. Gender differences were noted: Pre-MetS was more prevalent in females (66.92%) compared to males (59.48%). Central obesity was significantly higher among females (86.54%) than males (59.48%). The study also emphasized the socio-cultural factors influencing CMD prevalence, highlighting the role of dietary transitions and gendered health behavior.

## 4. Discussion

This systematic review reveals significant ethnic and regional variability in cardiometabolic risk across urban South Asian populations, particularly in India and the Eastern Indian state of Odisha. While urbanization is universally linked with higher cardiometabolic disease (CMD) burden, the degree and pattern of prevalence vary considerably based on socio-cultural, economic, and geographic contexts.

### 4.1 Urbanization and Lifestyle Transitions

The evidence consistently shows that urban environments are associated with elevated risk for CMDs due to lifestyle transitions such as reduced physical activity, increased intake of processed foods, and sedentary occupations [7,2]. The reviewed studies indicate a gradient of increasing CMD risk along the rural–urban continuum, where urban-middle-class populations exhibit the highest rates of obesity, hypertension, and diabetes [3]. These patterns reflect broader epidemiological shifts tied to rapid economic growth, globalization of food markets, and technological changes that reduce physical exertion in daily life.

### 4.2 Regional Disparities within Urban India

Within urban India, marked regional disparities in CMD prevalence were observed. Gupta et al. found that urban populations in western and northern India exhibit higher CMD risks than those in eastern regions [4]. This east-west gradient may reflect underlying differences in socio-economic development, dietary habits, caste/ethnic composition, and access to healthcare. For example, the relatively lower prevalence in eastern states like Odisha may be influenced by dietary patterns (e.g., lower fat consumption), lower urban density, or delayed exposure to globalized diets. However, as Odisha continues to urbanize, recent studies suggest a rising trend in metabolic syndrome and obesity, particularly among middle-aged and elderly adults [5].

### 4.3 Ethnic and Cultural Determinants

Ethnic and caste-based stratification, although underexplored, plays a crucial role in shaping CMD risk. South Asia’s complex socio-cultural structure—including caste, tribal affiliation, and religious practices—affects diet, physical activity, occupation, and healthcare-seeking behavior. While few studies disaggregate data by caste or tribal status, the higher prevalence of CMDs among urban females in Odisha may reflect gendered expectations and limitations in mobility or health access, especially in conservative or low-literacy communities [6]. The increased central obesity among women also aligns with cultural norms that associate a fuller figure with well-being and prosperity in some ethnic contexts [2].

### 4.4 Gender as an Intersecting Variable

Gender emerges as a consistent axis of disparity, particularly in eastern India. As highlighted by both Yasmin and Mohan et al., women exhibit a higher prevalence of central obesity and pre-metabolic syndrome [6,3]. This gender gap is likely shaped by reduced opportunities for physical activity, unequal access to nutritious food, caregiving burdens, and social taboos around women engaging in public exercise. These findings align with anthropological evidence suggesting that urban South Asian women’s health behaviors are constrained by socio-cultural expectations and limited autonomy, particularly in low-income households [4].

### 4.5 District-Level Insights: The Case of Kalahandi

The study conducted in urban Kalahandi offers a valuable lens into district-level cardiometabolic variation. Despite being a socio-economically marginalized district, Kalahandi’s urban populations show a high burden of CMDs. The finding that more than 63% of participants fall into the pre-MetS category underscores the silent buildup of risk before clinical disease manifestation [6]. The high prevalence among young adults (20–30 years) further suggests early exposure to urban stressors, dietary change, and sedentary behavior. This district-specific evidence reinforces the need for sub-state surveillance systems and culturally informed interventions, particularly as Tier 3 towns in India undergo rapid lifestyle transitions.

### 4.6 Policy Implications

Given the significant ethnic and regional heterogeneity observed, a “one-size-fits-all” approach to cardiometabolic health interventions in South Asia is unlikely to be effective. Instead, targeted strategies—such as promoting physical activity in culturally acceptable ways, strengthening primary healthcare at the municipal level, and incorporating traditional dietary wisdom—may yield better outcomes. Public health planning must also consider caste and tribal identity, gender norms, and urbanization level when designing awareness campaigns or screening programs.

### 4.7 Limitations

This review is limited by the availability of region- and ethnicity-specific data. Few studies provided granular breakdowns by caste, religion, or tribal status, and some relied on cross-sectional designs that preclude causal inference. The exclusion of non-English literature and the variation in CMD definitions across studies may also affect the comparability of findings. Nevertheless, the consistent trends observed strengthen the case for more localized, equity-focused cardiometabolic surveillance.

## 5. Conclusion

This systematic review highlights the significant ethnic and regional disparities in cardiometabolic risk across urban South Asian populations. The findings underscore the profound impact of urbanization, socioeconomic transitions, and culturally embedded behaviors on the rising burden of metabolic syndrome, diabetes, hypertension, and obesity in the region. While urbanization universally intensifies cardiometabolic risk, the degree and nature of this impact vary substantially across countries, states, and even districts—shaped by ethnicity, gender norms, caste dynamics, and access to healthcare.

In India, the gradient of risk from rural to urban areas is clear, yet even within urban centers, disparities persist based on geographic region and social identity. The evidence from Odisha, and specifically Kalahandi district, illustrates how socioeconomically marginalized regions are not exempt from CMD burden—in fact, they may experience a silent epidemic of pre-metabolic syndromes driven by uneven development, nutritional transition, and public health neglect.

These insights call for a paradigm shift from generalized national strategies to localized, culturally contextualized public health interventions. Addressing cardiometabolic risk in South Asia requires not only biomedical screening but also anthropological and sociological engagement with the communities at risk. Future research should prioritize disaggregated, intersectional data and explore the structural determinants that mediate disease vulnerability in urban South Asia.

## Data Availability

All data analyzed in this systematic review were extracted from previously published studies
available in public and subscription-based academic databases including PubMed, Google
Scholar, Scopus etc. These sources are cited in the references. No new data were generated in
this study

## Declaration by Authors

### Ethical Approval

**NA**

## Acknowledgement

**None**

